# Modeling the Spread of Misfolded Proteins in Alzheimer’s Disease using Higher-Order Simplicial Complex Contagion

**DOI:** 10.1101/2025.02.01.25321521

**Authors:** Marcin Wardynski, Iacopo Iacopini, Giovanni Petri, Vito Latora, Alessandro Crimi

**Affiliations:** Faculty of Computer Science, AGH University of Krakow., Krakow, Poland; Network Science Institute, Northeastern University London, London UK; School of Mathematical Sciences Queen Mary, University of London, London, UK

## Abstract

Neurodegenerative diseases are characterized by complex proteins misfolded that propagate within the brain. For instance, current findings highlight the role of 2 specific misfolded proteins in Alzheimer which are believed to spread using brain fibers as highways. Previous studies investigated such spreading by simulation models or machine learning-based predictors which adopt the brain connectome as the underlying spreading network. However, the structural connectome by construction only describes pairwise connections between nodes in a graph. High-order interaction complex networks offer significant advantages over normal graphs because they can capture interactions that go beyond simple pairwise relationships. Protein misfolding and aggregation often involve cooperative behaviors or group dynamics that normal graphs, with their focus on individual edges, cannot adequately represent. The non-linear and multiscale nature of protein misfolding might be better suited to a richer representation of higher-order models. In this study we investigate whether higher-order networks can provide improved fits and explanatory power in this context. More specifically, we employ a simplicial complex contagion model for amyloid beta to predict protein misfolding spread. The simplicial contagion complex produced a mean reconstruction error of 0.030 for Alzheimer’s patients regarding the predicted protein deposition across all brain region in a 2-year horizon and other results, outperforming previous studies, especially for cases in which the misfolded protein were non-increasing steadily. Despite the limited time span, this study highlights the potential of combining advanced network analysis to capture the intricate dynamics of protein aggregation across neural networks.

**Clinical relevance:** This study highlights the potential of high-order networks to improve predictions of misfolded protein spread in Alzheimer’s, offering better insight into protein aggregation dynamics.

## I. Introduction

Protein misfolding represents a critical pathological mechanism in neurodegenerative disorders, with progressive neural system dysfunction fundamentally altering network connectivity [1]. Traditional models have limitations in capturing the complex spatial and temporal dynamics of protein propagation [2]. Unlike earlier models that viewed protein aggregation as a localized cellular event, current research demonstrates a complex intercellular transmission mechanism through which misfolded proteins propagate through neuronal networks via multiple potential routes [2]. In recent years, Diffusion-weighted imaging (DWI) showed information on the anatomical connectivity inside the brain, polarizing water molecules to detect their diffusion in tissues to identify white matter tracts through tractography. The structural connectome [3] represents the brain network using regions, provided by an anatomical atlas [4], as nodes, and white fibers as weighted edges, exploiting graph theory to obtain detailed measures on biological processes flowing within. Indeed, the accepted paradigm in Alzheimer’s disease (AD) suggests that diffusion patterns revolve mostly around two misfolded proteins, namely *Amyloid β*− (*Aβ*) and *tau* (*τ*), across neuronal pathways [5]. Through radiopharmaceutical tracers, it is possible to obtain positron emission tomography (PET) images to measure the concentrations of proteins to estimate the prediction error of this guess and to consequently improve its undertaking [6]. Indeed, computational models have been proposed, those have been either inspired by spreading mechanism similar to the diffusion of heat [7], or similar to epidemic spreading [8]. In parallel, machine learning approaches using autoregressors and graph convolutional networks achieved comparable results without the use of simulations [9].

The Braak staging is one of the oldest systems used to describe the spread in a predictable pattern across specific brain regions as the disease progresses. This staging provides insight into the disease’s severity, and helps link the observed pathology to clinical symptoms. However, not all patients follow the exact sequence outlined by the Braak staging [5]. Moreover, their spatio-temporal trajectories are discordant, *Aβ* might even slow down at later stages, as opposed to *τ*, which continues to increase steadily for the remaining course of AD [6]. In this view, the disease has mostly 2 stages. One initial with the diffusion of *Aβ* which does not correlate well with symptoms, and one more advanced with the spreading of *τ* filaments which correlates better with symptoms [6], as shown in Figure 2. This conceptualization allowed to define an AD biological model with physiological changes [10]. Indeed, *τ* filaments are considered at the moment more indicative for diagnosis. Nevertheless, it can be hypothesized that, to stop disease progression, it is more relevant to act with therapies such as trans-cranial stimulation [11] or pharmaceutical [12] at early stage when only the *Aβ* are present. Therefore we focus on this stage as we are more interested in prediction at an early stage.

Indeed, our analysis showed that *Aβ* behaves similarly to a sigmoidal temporal evolution, where after a steep increase, the overall deposition and aggreagation of proteins does not increase, as shown in other studies [6]. Unfortunately, it is not as straightforward to understand which patients are in the steep increase phase or in the plateaux. To further complicate this picture, some patients might show only mild cognitive impairment, which may remain so, without a complete conversion to AD patients. In this study, we take into account this heterogeneity, evaluating how predictor models react while attempting to predict future states of patients whether they are among those with increasing protein deposition or in plateaux. We do this by expanding the previous comparison between spreading models and machine learning predictors [9] using more advanced complex networks relationships (as depicted in Figure 1). More in detail, this study aims to(i)evaluate a spreading model using simplicial complex, and (ii) quantify protein misfolding spread probabilities across heterogeneous subjective patterns. By leveraging recent advances in network science, machine learning, and computational neurobiology, we propose a novel framework for understanding and predicting neurodegenerative disease progression. Using this approach, we predict the deposition of misfolded protein with a horizon of 2 years.

**Fig. 1.**
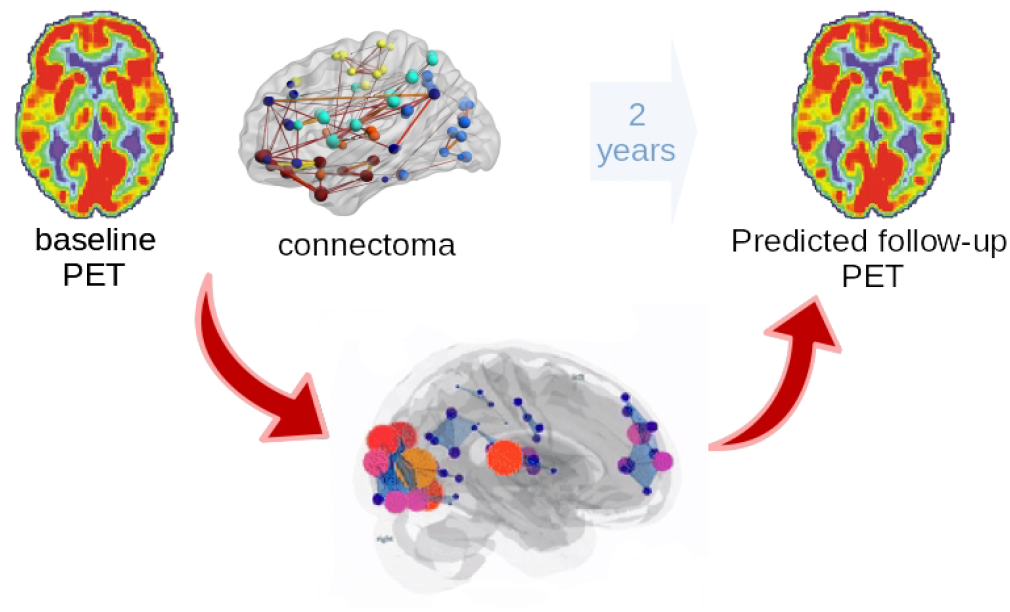
The high-level pipeline where a baseline PET for protein deposition and a structural connectivity matrix is used to define either a high-order spreading model, predicting PET deposition after 2 years.

## II. Methods

### A. Data and preprocessing

We used a subset of the ADNI defined by the subjects and patients for which all the required modalities where available (DWI, T1-weighted, and PET). Ethical approval and patient consent statement were not necessary as previously acquired by ADNI, and information about the approval are available on their website. The resulting demographics for the selected subjects are the following: AD subjects (age: 76.5 ± 7.4 years), control (CN) (age 77.0 ±5.1 years), mild cognitive impaired (MCI) (age: 75.34 ± 5.93); with relatively uniform sex distribution (52% male and 48% female subjects). Data were processed as in [9] and accessible on Figshare ^1^. Briefly, the pre-processing steps were the following: The DWI data were skulls-stripped, denoised and eddy current corrected, and structural connectivity was obtained by using the DiPy library and the Automated Anatomical Labeling 3 (AAL3) [13], leading to 166 ROIs, with an average of 22.9 edges between ROIs. For the PET data, we performed motion correction through a co-registration of frames, then we computerd the average of these frames, over time, using fslmaths, then registered to a reference volume of AAL3 with skull, and skull stripped afterwards. We further normalized PET images of each subject by the maximum value of the image in the range [0, 1], and computed the average regional concentrations for each image following the ROIs of AAL3. We again averaged for each category (AD, MCI and control subjects) and applied a z-score normalization against baseline PET concentrations for CN subjects as in [7]. Then, z-scores were remapped into a [0, 1] interval using a standard logistic function. The predicted protein concentration values are tested with ground-truth values given in PET follow-up scans after 2 years.

Our experiments are carried out taking into account the different clinical statuses: CN, MCI, and AD. Moreover, following the insight of the sigmoidal behavior of the *Aβ* deposition as shown in Figure 2, we take into account further stratification. Practically, we investigate further how the predictions result separately for subjects where the protein deposition is increasing within the considered interval or not. Those values are reported in the tables in Section Results and Discussions. In our study, the subjects that are considered non-increasing are defined as

**Fig. 2.**
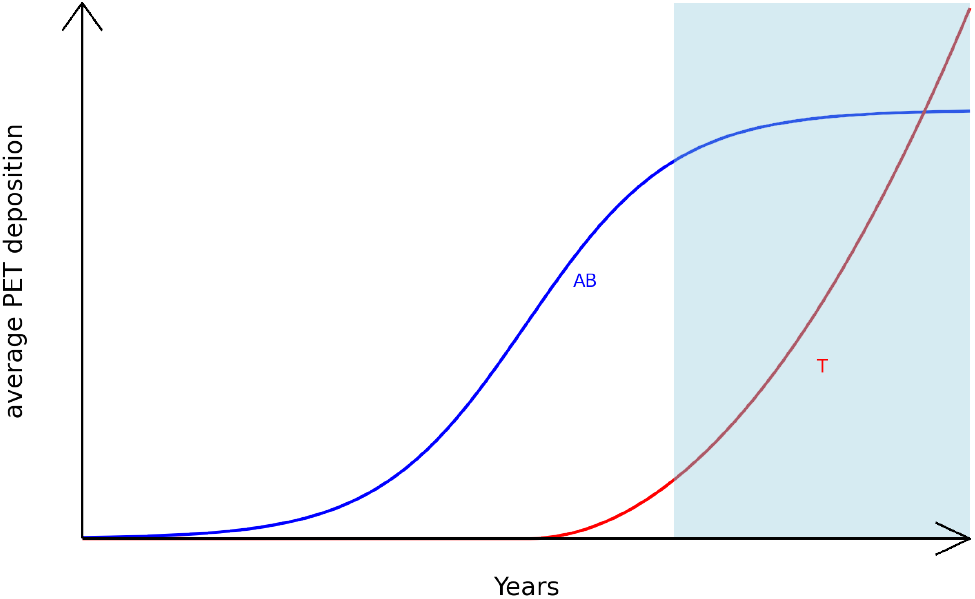
Simplistic view of the spreading of misfolded proteins associated to Alzheimer’s. Initially the accumulation of *Aβ* with little relationship with symptoms and believed to saturate at some points, then exponential spreading of *τ* -filaments with more reliable relationships with cognitive deficits marked by the darker area.

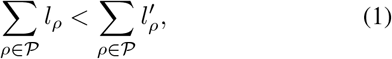

where *ρ* stands for a single region, 𝒫 is the set of all considered regions, *l*_*ρ*_ and 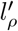are *Aβ* concentrations in specific regions when measured for the first and second time respectively.

### B. Simplicial Complex Contagion

Extending traditional network representations, we adopt the simplicial complex approach pioneered by Iacopini *et al*. to model protein misfolding propagation. Unlike traditional graph models that capture only pairwise interactions, simplicial complexes enable the representation of higher-order interactions and dependencies in biological networks. A simplicial complex 𝒦 is defined as a collection of simplices *σ* ∈ 𝒦, where a simplex is a generalized concept of an edge representing k-dimensional interactions:

- 0-simplex: Individual nodes (brain regions)
- 1-simplex: Edges (pairwise interactions)
- 2-simplex: Triangles (three-way interactions)
- *k*-simplex: Higher-dimensional interactions

This extends traditional graph representations by incorporating higher-order interactions as Δ^*k*^ = {(*v*_0_, …, *v*_*k*_) : *v*_*i*_ ∈*V*, 0≤ *i* ≤*k*}, allowing also multidimensional modeling of protein network propagation [14]. Specifically, the spread of misfolded protein across networked brain regions is mathematically modeled using the Susceptible-Infected-Susceptible (SIS) compartmental framework, traditionally introduced in the context of infectius diseases. Here, the SIS model describes the transition from healthy regions (S) to regions containing a non-zero concentration of misfolded proteins (I). Transition between compartments are governed by rates of “infection” (from S to I, via contacts with I nodes) and “recovery” (from I to S). We consider the simplicial contagion model which has been introduced to model behavioral contagion in social networks [14]. It is adapted here as following: Nodes represent brain regions linked by fiber bundles, with initial contagion probabilities based on baseline *Aβ* levels from PET imaging. Edges denote brain region connections from the connectome, ignoring weights. Simplices capture protein interactions within cliques, such as cellular transfers, extracellular vesicle pathways, and neuronal connectivity. For each subject, a separate graph is constructed according to the aforementioned rules from the diffusion MRI data. Moreover, the framework is restricted to 2-simplices of 𝒦 (triangles) as in Figure 3, since our implementation currently does not include higher-order simplices. However, it can be easily extended to *k*-simplices. The dynamics of the system evolves according to a microscopic Markov chain approach (MMCA) [15]. At the node level, the concentration of misfolded protein at time *t* is calculated based on levels at time *t* − 1, using the following equation:

**Fig. 3.**
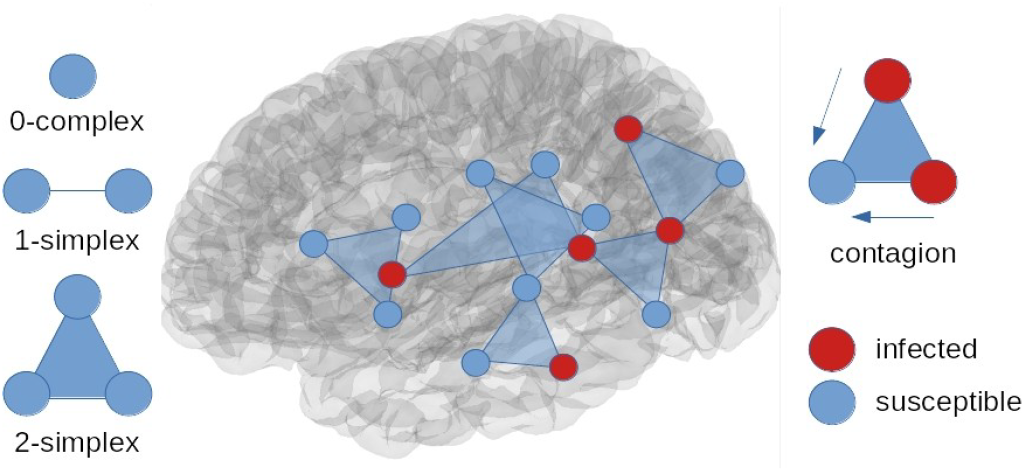
High-order spreading of protein uses underlying d-dimensional group interactions as depicted on the left, all over the brain as shown in the middle, and those are reflected in high-order contagions as shown on the right.

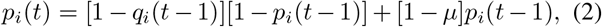

with

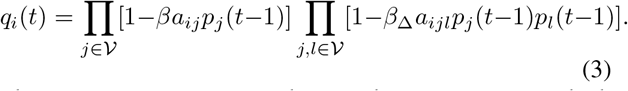

The parameter *µ* in Eq. (2) denotes the recovery rate, which is node-independent and constant in time. Contrarily, the parameters *β* and *β*_Δ_ that appear in Eq. (3) represent the transmission rates for 1- and 2-simplices, respectively. The structure is encoded into the elements of the adjacency matrices *a*_*ij*_ and *a*_*ijl*_ that return 1 if the 1-simplex *ij*, or the 2-simplex *ijl*, exists, and 0 otherwise.

Generally, misfolded protein concentrations do not decrease—the transition from I to S via *µ* should not be allowed. Nevertheless, here the recovery rate *µ* is introduced to model confounding factors such as changes in diets and sleeping habits which could relatively reduce protein concentrations [16], [17].

We fit the model using Optuna based on tree-structured Parzen stimator with Gaussian mixture model over multiple simulations, each consisting of 50 steps throughout the dataset. Before fitting, infectivity parameters are rescaled in terms of ⟨*k*⟩ and ⟨*k*_Δ_⟩, the average number of incident 1- and 2-simplices per node, respectively. This leads to the new variables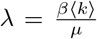and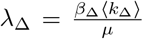. The resulting best-fitting parameters are *λ* = 0.20, *λ*_Δ_ = 2.21, and *µ* = 0.0003. Such a small *µ* value justifies the a priori assumption, given its limited impact compared to the infection parameters.

## III. Results and Discussion

The differentiation between subjects with increasing and non-increasing protein deposition using the equation 1, led the identification of subjects in all clinical groups. Those are reported in Table I. This shows that less than half of the subjects has non-increasing deposition using equation 1. This large fraction might be related to the fact we are limited by the available data. Namely, there is only one interval of 2 years between baseline and follow-up PET, while the entire evolution, as ideally depicted in Figure 2, might span 50-60 years. Despite this, our model demonstrates significant improvements in tracking protein misfolding patterns. More precisely, we compared our results to a previous model based on MAR which was already showing superior performance compared to other state of the art models [9] as network diffusion model [7] and epidemic spreading [8]. The rationale behind why a simplical contagion outperforms an autoregressive model and also other spreading models based on graphs rather than hypergraphs, can be given by the fact that a simplicial complex can capture the non-linearity of protein spreading given by a clique rather than just an edge [14]. A MAR model can be used in various flavors, for example, knowing that a subject is an Alzheimer patient, we can use a model trained on data of the same type to reach a better *Aβ* concentration forecast. In this case, the results reported in Table II highlights the MAR model as performing better than the proposed model. However, without knowing the diagnosis enforces the use of the general purpose MAR trained on all the data. We would consider this a more realistic clinical scenario in which the proposed model performs better as reported in bold in Table III and IV.

**TABLE I.** Distribution of cases across categories by increasing or non-increasing protein deposition within 2 years.

| Category | AD | LMCI | MCI | EMCI | CN |
| --- | --- | --- | --- | --- | --- |
| Total | 24 | 47 | 30 | 60 | 51 |
| Non-Increasing | 16 | 20 | 9 | 29 | 26 |

**TABLE II.** Comparative Model Performance for Diagnosed Subject.

| Category | MAR |  | SCM |  |
| --- | --- | --- | --- | --- |
| | MSE$\pm\sigma$ | PCC$\pm\sigma$ | MSE$\pm\sigma$ | PCC$\pm\sigma$ |
| AD | **.0027 ± .00** | .9662 ± .02 | .0030 ± .00 | .9662 ± 0.02 |
| LMCI | .0066 ± .02 | .9231 ± .18 | **.0064 ± .02** | .9225 ± 0.18 |
| MCI | **.0105 ± .01** | .7684 ± .28 | .0107 ± .01 | .7677 ± 0.28 |
| EMCI | .0065 ± .02 | .9171 ± .20 | **.0064 ± .02** | .9163 ± 0.20 |
| CN | .0044 ± .01 | .9457 ± .10 | **.0044 ± .01** | .9453 ± 0.10 |
| ALL | .0062 ± .02 | .9097 ± .19 | **.0061 ± .02** | .9093 ± 0.19 |

**TABLE III.** Comparative Model Performance for Undiagnosed Subject.

| Category | MAR (I w. struct) |  | SCM |  |
| --- | --- | --- | --- | --- |
| | MSE$\pm\sigma$ | PCC$\pm\sigma$ | MSE$\pm\sigma$ | PCC$\pm\sigma$ |
| AD | .0034 ± .00 | .9664 ± .02 | **.0030 ± .00** | .9662 ± .02 |
| LMCI | .0066 ± .02 | .9232 ± .18 | **.0064 ± .02** | .9225 ± .18 |
| MCI | **.0105 ± .01** | .7684 ± .28 | .0107 ± .01 | .7677 ± .28 |
| EMCI | .0065 ± .02 | .9167 ± .20 | **.0064 ± .02** | .9163 ± .20 |
| CN | .0044 ± .01 | .9457 ± .10 | **.0044 ± .01** | .9453 ± .10 |
| ALL | .0062 ± .02 | .9097 ± .19 | **.0061 ± .02** | .9093 ± .19 |

**TABLE IV.** Model Performance for Non-Increasing *Aβ* Concentration.

| Category | MAR (I w. struct) |  | SCM |  |
| --- | --- | --- | --- | --- |
| | MSE$\pm\sigma$ | PCC$\pm\sigma$ | MSE$\pm\sigma$ | PCC$\pm\sigma$ |
| AD | .0042 ± .00 | .9667 ± .02 | **.0035 ± .00** | .9663 ± .02 |
| LMCI | .0113 ± .04 | .8920 ± .26 | **.0103 ± .03** | .8909 ± .26 |
| MCI | .0068 ± .01 | .9435 ± .05 | **.0059 ± .01** | .9428 ± .05 |
| EMCI | .0099 ± .03 | .9164 ± .20 | **.0091 ± .03** | .9160 ± .20 |
| CN | .0029 ± .01 | .9651 ± .02 | **.0025 ± .00** | .9649 ± .02 |
| ALL | .0072 ± .02 | .9347 ± .16 | **.0064 ± .02** | .9342 ± .16 |

## IV. Conclusion

The proposed simplicial graph model provides a sophisticated computational framework for understanding protein misfolding propagation. The simplicial contagion approach shows higher accuracy compared to the model that had previously achieved better performance, offering potential insight into the mechanisms of neurodegenerative diseases. This is particularly relevant to conditions like Alzheimer’s and Parkinson’s disease which are critical to understand stratification of patients [18]. Future works include repeating those analysis with longer horizon in the prediction and also investigating *τ* filament depositions, or even expand the model with hypergraph convolutional networks [19].

## Data Availability

All data used are available online at https://adni.loni.usc.edu/

## Footnotes

1 https://figshare.com/articles/dataset/Pre-processed_data_for_the_study_Gherardini_et_al_/22645357

## References

[1] W. Seeley et al., “Neurodegenerative diseases target large-scale human brain networks,” Neuron, vol. 62, no. 1, pp. 42–52, 2009.

[2] J. Vogel et al., “Connectome-based modelling of neurodegenerative diseases: towards precision medicine and mechanistic insight,” Nature Reviews Neuroscience, vol. 24, no. 10, pp. 620–639, 2023.

[3] P. Hagmann et al., “Mapping the structural core of human cerebral cortex,” PLOS Biology, vol. 6, no. 7, pp. 1–15, 07 2008.

[4] S. B. Eickhoff, B. T. T. Yeo, and S. Genon, “Imaging-based parcellations of the human brain,” Nature Reviews Neuroscience, vol. 19, no. 11, pp. 672–686, Nov 2018.

[5] H. Braak and E. Braak, “Staging of Alzheimer’s disease-related neurofibrillary changes,” Neurobiology of Aging, vol. 16, no. 3, pp. 271–278, 1995.

[6] M. S. Baek et al., “Temporal trajectories of in vivo tau and amyloid-β accumulation in Alzheimer’s disease,” European Journal of Nuclear Medicine and Molecular Imaging, vol. 47, pp. 2879–2886, 2020.

[7] A. Raj, E. LoCastro, A. Kuceyeski, D. Tosun, N. Relkin, and M. Weiner, “Network diffusion model of progression predicts longitudinal patterns of atrophy and metabolism in Alzheimer’s disease,” Cell Reports, vol. 10, no. 3, pp. 359–369, 2015.

[8] Y. Iturria-Medina, R. C. Sotero, P. J. Toussaint, A. C. Evans,, and the Alzheimer’s Disease Neuroimaging Initiative, “Epidemic spreading model to characterize misfolded proteins propagation in aging and associated neurodegenerative disorders,” PLOS Computational Biology, vol. 10, no. 11, pp. 1–16, 11 2014.

[9] L. Gherardini et al., “Prediction of misfolded proteins spreading in Alzheimer’s disease using machine learning and spreading models,” Cereb Cortex, vol. 33, no. 24, p. 11471–11485, 2023.

[10] C. R. Jack et al., “A/T/N: An unbiased descriptive classification scheme for Alzheimer disease biomarkers,” Neurology, vol. 87, no. 5, pp. 539–547, 2016.

[11] L. Pini, F. B. Pizzini, I. Boscolo-Galazzo, C. Ferrari, S. Galluzzi, M. Cotelli, E. Gobbi, A. Cattaneo, et al., “Brain network modulation in Alzheimer’s and frontotemporal dementia with transcranial electrical stimulation,” Neurobiology of Aging, vol. 111, pp. 24–34, 2022.

[12] L. Pini, S. Lista, A. Griffa, G. Alalli, and B. P. Imbimbo, “Can brain network connectivity facilitate the clinical development of disease-modifying anti-Alzheimer drugs?” Brain Communications, 2025.

[13] E. T. Rolls, C.-C. Huang, C.-P. Lin, J. Feng, and M. Joliot, “Automated anatomical labelling atlas 3,” NeuroImage, vol. 206, p. 116189, 2020.

[14] I. Iacopini et al., “Simplicial models of social contagion,” Nature Communications, vol. 10, no. 2485, pp. 193–200, 2019.

[15] I. Iacopini, “Modelling the social dynamics of contagion and discovery using dynamical processes on complex networks,” Ph.D. dissertation, Queen Mary University of London, 2021. [Online]. Available: https://iaciac.github.io/publication/phd-thesis/

[16] L. Xie, H. Kang, Q. Xu, M. J. Chen, Y. Liao, M. Thiyagarajan, J. O’Donnell, D. J. Christensen, C. Nicholson, J. J. Iliff et al., “Sleep drives metabolite clearance from the adult brain,” Science, vol. 342, no. 6156, pp. 373–377, 2013.

[17] O. Lazarov, M. P. Mattson, D. A. Peterson, S. W. Pimplikar, and H. van Praag, “When neurogenesis encounters aging and disease,” Trends in neurosciences, vol. 33, no. 12, pp. 569–579, 2010.

[18] C. Abdelnour, F. Agosta, M. Bozzali, B. Fougère, A. Iwata, R. Nilforooshan, L. T. Takada, F. Viñuela, and M. Traber, “Perspectives and challenges in patient stratification in Alzheimer’s disease,” Alzheimer’s research & therapy, vol. 14, no. 1, p. 112, 2022.

[19] Z. Zhang, Y. Feng, S. Ying, and Y. Gao, “Deep hypergraph structure learning,” arXiv preprint arXiv:2208.12547, 2022.

